# SARS-CoV-2 spike-binding antibody longevity and protection from re-infection with antigenically similar SARS-CoV-2 variants

**DOI:** 10.1101/2022.03.28.22273068

**Authors:** John Kubale, Charles Gleason, Juan Manuel Carreño, Komal Srivastava, PARIS Study Team, Aubree Gordon, Florian Krammer, Viviana Simon

## Abstract

The PARIS (**P**rotection **A**ssociated with **R**apid **I**mmunity to **S**ARS-CoV-2) cohort follows health care workers with and without documented coronavirus disease 2019 (COVID-19) since April 2020. We report our findings regarding SARS-CoV-2 spike binding antibody stability and protection from infection in the pre-variant era. We analyzed data from 400 healthcare workers (150 seropositive and 250 seronegative at enrollment) for a median of 84 days. The SARS-CoV-2 spike binding antibody titers were highly variable with antibody levels decreasing over the first three months, followed by a relative stabilization. We found that both more advanced age (>40 years) and female sex were associated with higher antibody levels (1.6-fold and 1.4-fold increases, respectively). Only six percent of the initially seropositive participants “seroreverted”. We documented a total of 11 new SARS-CoV-2 infections (ten naïve participants, one previously infected participant without detectable antibodies, p<0.01) indicating that spike antibodies limit the risk of re-infection. These observations, however, only apply to SARS-CoV-2 variants antigenically similar to the ancestral SARS-CoV-2 ones. In conclusion, SARS-CoV-2 antibody titers mounted upon infection are stable over several months in most people and provide protection from infection with antigenically similar viruses.

**summary:** The levels of SARS-CoV-2 spike binding antibodies mounted upon infection with ancestral SARS-CoV-2 variants are highly variable, stabilize at an individual level after three months and provide protection from infection with homologous virus.

## Introduction

Severe acute respiratory syndrome coronavirus 2 (SARS-CoV-2) caused the coronavirus disease 2019 (COVID-19) pandemic with over 430 million infections (WHO dashboard, February 25^th^ 2022) since it emerged in late 2019 (1, 2). In the vast majority of individuals, infection with SARS-CoV-2 leads to the induction of a specific adaptive immune response including spike binding as well as neutralizing antibodies (3, 4). Indeed, we found that over 90% of individuals infected during the first wave in New York City (NYC) had robust antibody titers as measured using an enzyme-linked immunosorbent assay (ELISA, >30,000 cross-sectional measurements) (4, 5). The durability and protective effect of such antibody responses remains a topic of active investigation even as we move into the third year of the pandemic. An initial study (6) reported fast waning of SARS-CoV-2 binding antibodies but others report spike binding IgG antibodies being detectable months after infection (7-9).

The first SARS-CoV-2 infection in New York State was officially detected at the Mount Sinai Health System in NYC on February 29^th^, 2020 although SARS-CoV-2 had likely been introduced to the local communities weeks to months earlier (10, 11). Indeed, the New York metropolitan area emerged as one of the early COVID-19 epicenters in the US. This initial COVID-19 wave was exponential in growth and nearly overwhelmed our local health care systems due to the high number of patients with severe COVID-19 manifestations resulting in infection fatality rates ranging between 1 and 1.5% (11, 12). It was at that point in time (April 2020) that we started enrollment for the PARIS (**P**rotection **A**ssociated with **R**apid **I**mmunity to **S**ARS-CoV-2) cohort to follow health care workers (HCWs) of the Mount Sinai Health System with and without documented COVID-19 over time. Full-length spike binding IgG antibody titers were measured every two to four weeks using a sensitive and specific quantitative ELISA (13). In addition, data on potential exposures as well as clinical signs and symptoms suggestive of SARS-CoV-2 infection were collected at the same time intervals.

Here we report our findings regarding the kinetics of SARS-CoV-2 spike binding IgG antibody titers over time and the protection from reinfection in a high-risk work environment.

## Material and Methods

### Description of the PARIS cohort

The PARIS study enrolled healthcare workers with and without prior SARS-CoV-2 infection to study the durability and effectiveness of the immune response to SARS-CoV-2. A total of 501 participants were enrolled between April 2020 and August 2021. The study protocol was reviewed and approved by the Mount Sinai Hospital Institutional Review Board (IRB-20-03374). All participants provided written informed consent. Samples were coded prior to processing and testing. Blood was collected at 2-4 week intervals regardless of the serostatus at enrollment. For this analysis, the cohort was restricted to 400 participants enrolled prior to SARS-CoV-2 vaccination with, at least, four weeks of follow-up or two study visits prior to vaccination. At the time of enrollment, 150/400 participants were seropositive for SARS-CoV-2 spike binding antibodies while 250/400 were seronegative. Most participants had no known immunosuppressive conditions/co-morbidities. We used the data from 2,106 distinct study visits from these 400 participants to evaluate risk of SARS-CoV-2 infection and seroreversion. From this dataset, we selected a subset of 137 participants with known dates of COVID-19 (symptom onset, positive SARS-CoV-2 nucleic acid amplification test (NAAT), or positive SARS-CoV-2 antibody test results) and, at least, two pre-vaccine study visits with SARS-CoV-2 antibody measurements. The data of 813 distinct study visits from these 137 seropositive participants provide the basis for the modeling SARS-CoV-2 spike-binding IgG antibody durability.

### Identification of new SARS-CoV-2 infections in PARIS

One of the 11 participants who were infected during the observation period was diagnosed as part of the study using viral diagnostic NAAT while nine participants tested positive for SARS-CoV-2 outside of the Mount Sinai Health System. One asymptomatic infection was identified by seroconversion (from negative to a titer of 1:400).

### SARS-CoV-2 full-length spike binding antibody measurements

Antibody titers were determined using a two-step ELISA protocol (13) in which sera are screened at a single dilution (1:50) for IgG against the recombinant receptor binding domain (RBD) of the spike protein from SARS-CoV-2, followed by detection of antibodies against the full-length spike protein. End-point titers were determined by serially diluting sera (from 1:80 to 1:102,400). Briefly, 96-well microtitre plates (Thermo Fisher) were coated with 50 μl/well of recombinant RBD (2 μg/ml) overnight at 4 °C. Plates were washed three times with phosphate-buffered saline (PBS; Gibco) supplemented with 0.1% Tween-20 (PBS-T; Fisher Scientific) using an automatic plate washer (BioTek). Plates were then blocked with PBS-T containing 3% milk powder (American Bio) for 1 hour. Sera was heat-inactivated and serially diluted (3-fold) in PBS-T 1%-milk powder, starting at 1:50 initial dilution for RBD ELISA and at 1:80 dilution for full-length spike ELISA. Samples were added to the plates and incubated for 2-hours. Plates were washed three times with PBS-T and 50 μl/well of anti-human IgG (Fab-specific) horseradish peroxidase antibody (produced in goat; Sigma, A0293) diluted to 1:3,000 in PBS-T, 1% milk powder, were added to each well. After 1-hour incubation at room temperature, plates were washed three times with PBS-T and 100 μl/well of SigmaFast o-phenylenediamine dihydrochloride (Sigma) were added for 10min, followed by addition of 50 μl/well of 3M hydrochloric acid (Thermo Fisher) to stop the reaction. Optical density was measured at a wavelength of 490 nm using a plate reader (BioTek). Endpoint titers, expressed as the last dilution before the signal dropped below an OD490nm of 0.15, were calculated in excel and data was plotted using GraphPad Prism 9.

### Assessment and modeling of SARS-CoV-2 Spike binding antibody durability over time

To assess how spike antibody titers changed over time we fit an additive mixed model using the mgcv package (version 1.8-36) for R (version 4.1.1). Participants were excluded from the model if they never developed a detectable titer (at least 1:80) during follow-up or if the data regarding when they were infected (illness onset, NAAT positive, or antibody positive date) was missing. Day 0 was defined as the reported symptom onset date or the date of positive SARS-CoV-2 diagnostic test.

Spike titers (ranging from 1:80 to 1:6,400) were transformed to the log2 scale so that a 1-unit increase corresponded to a doubling of antibody titer. Sex (female, male), age (<40 years, 40+ years), and baseline titer (<1:800, ≥1:800) were included as covariates in the model, along with a random intercept for participant ID to account for repeated measures. Finally, a penalized spline term was included to model antibody titer over time. We fit this model in two ways: 1) assuming that antibody decay occurred at the same rate over time regardless of baseline titer, 2) allowing antibody decay over time to vary by baseline titer. In the second model a distinct smoothing function was fit for each baseline titer group. Data is available upon request, and the code used for modeling is available on GitHub at https://github.com/jkubale/paris.

### Determination of the frequency of spike binding antibody seroreversion

Seropositive participants who initially had measurable spike binding antibodies but subsequently had spike binding antibody levels below the limit of detection (1:80) on two consecutive visits were defined as having seroreverted. We examined the probability of seroreversion over time for those with low (<1:800) and high (≥1:800) baseline titers by calculating the probability of survival (not seroreverting) via the Kaplan-Meier estimator.

### Assessment of protection against re-infection

We explored whether participants with a detectable antibody titer had a lower probability of incident SARS-CoV-2 infection. New SARS-CoV-2 infections were identified by positive NAAT or by SARS-CoV-2 antibody seroconversion. The participant immune status was based on the most recent spike binding antibody titer preceding the infection. Participants with detectable spike binding antibody titers were compared to those without detectable titers using Fisher’s Exact Test. All analyses were performed using R version 4.1.1.

## Results

We analyzed the spike binding IgG antibody levels of 400 PARIS participants with (N: 150) or without previous COVID-19 (N: 250) collected every two to four weeks for a median of 84 days (IQR: 55-169) from April 2020 to August 2021. The majority of participants were female (68%) with a median age of 35 years (range: 19-75; interquartile range [IQR]: 30-45). The demographics of the cohort are summarized in **Table 1**. Approximately a third of the participants self-reported as performing high-risk tasks as part of their work assignments. Most participants with spike binding antibodies at study enrollment (92.7%, 139/150) were infected during the first pandemic wave when NYC was one of the epicenters of the pandemic (March-May 2020). Of the remaining 11 participants, seven were infected in the summer and fall of 2020 prior to enrolling into PARIS and four participants did not recall having any symptoms suggestive of COVID-19. Two PARIS cohort datasets were used to analyze durability and effectiveness of serological responses (Protection Dataset, Antibody Durability Dataset, **Figure 1A, Table 1**).

**Table 1:** Characteristics of the PARIS participants included in the Protection Dataset (A; N=400) as well as in the Antibody Durability Dataset (B; N=137).

| **A: Protection Dataset** |  |  |  |
| --- | --- | --- | --- |
|  | **Total** | **Seronegative** | **Seropositive** |
| **Number of Participants** | 400 (100.0%) | 250 (62.5%) | 150 (37.5%) |
| **Sex** |  |  |  |
| Female | 273 (68.3%) | 175 (70.0%) | 98 (65.3%) |
| Male | 126 (31.5%) | 74 (29.6%) | 52 (34.7%) |
| Prefer Not to Say | 1 (0.3%) | 1 (0.4%) | 0 (0.0%) |
| **Age** |  |  |  |
| <40 | 250 (62.5%) | 162 (64.8%) | 88 (58.7%) |
| 40+ | 149 (37.3%) | 87 (34.8%) | 62 (41.3%) |
| Missing | 1 (0.3%) | 1 (0.4%) | 0 (0.0%) |
| **Days Enrolled (median, IQR)** | 84 (55-169.25) | 84 (55.25-174) | 84 (55-167) |
| **Seroreversion** | 9 (2.3%) | - | 9 (6.0%) |
| **B: Antibody Durability Dataset** |  |  |  |
|  | **Total** | **High baseline titer (800+)** | **Low baseline titer (<800)** |
| **Number of Participants** | 137 (100.0%) | 81 (59.1%) | 56 (40.9%) |
| **Sex** |  |  |  |
| Female | 89 (65.0%) | 59 (72.8%) | 30 (53.6%) |
| Male | 48 (35.0%) | 22 (27.2%) | 26 (46.4%) |
| **Age** |  |  |  |
| <40 | 80 (58.4%) | 37 (45.7%) | 43 (76.8%) |
| 40+ | 57 (41.6%) | 44 (54.3%) | 13 (23.2%) |
| **Days Enrolled (median, IQR)** | 84 (56-168) | 108 (56-192) | 69 (55-112) |
| **Days between SO<sup>b</sup> and Enrollment** | 154 (91-203) | 147 (63-196) | 182 (133-217) |
| **Baseline SARS-CoV-2 Antibody Titer<sup>c</sup>** | 1:730 (2.67) | 1:1419 (1.93) | 1:279(1.52) |
| **Seroreversion** | 8 (5.8%) | 0 (0.0%) | 8 (14.3%) |

<sup>a</sup> N (%)
<sup>b</sup> SO = symptom onset: median (IQR)
<sup>c</sup> Geometric mean (Standard Deviation)

**Figure 1:**
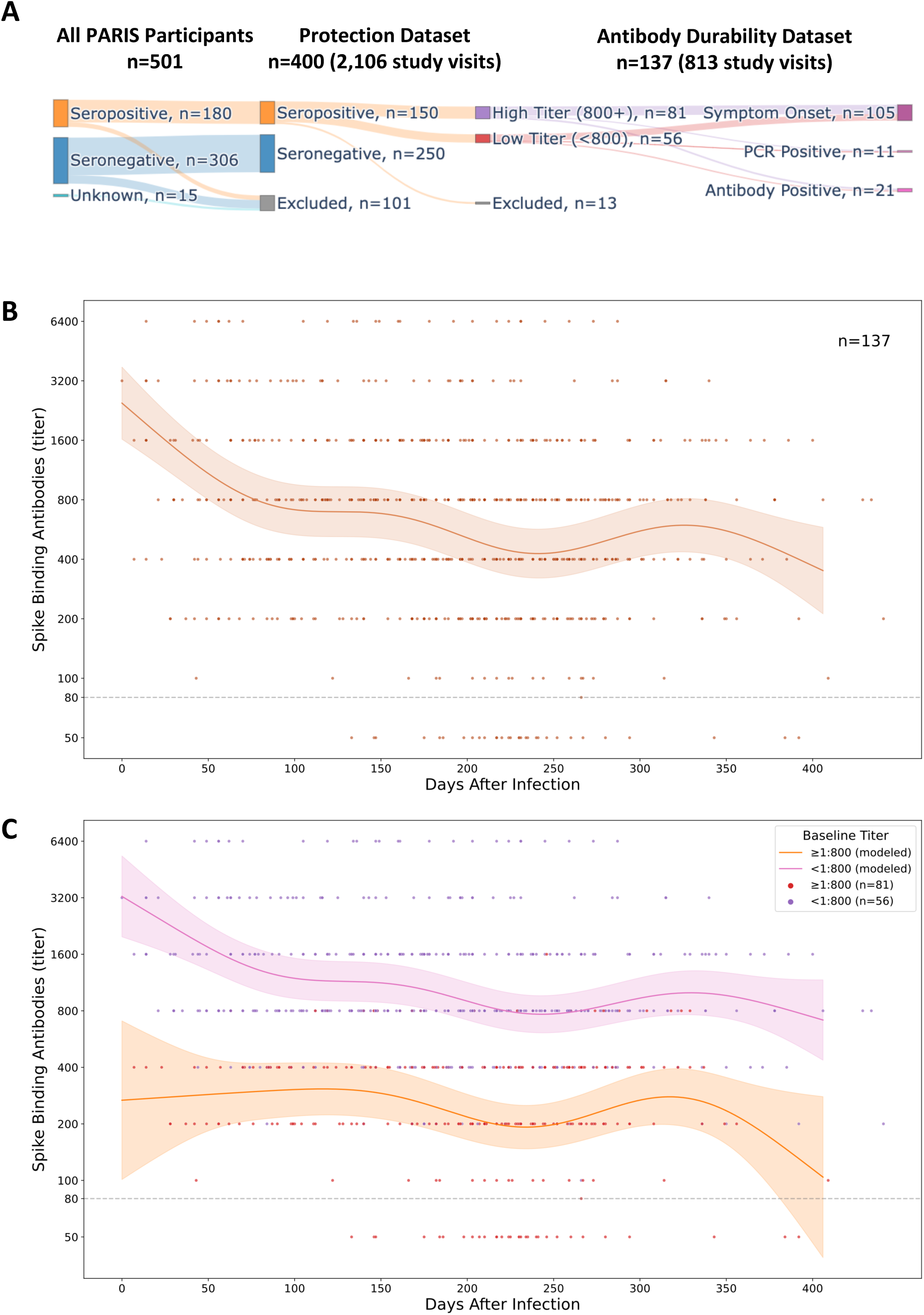
Modeling SARS-CoV-2 antibody durability in PARIS participants. A: Overview of the PARIS cohort datasets (protection, antibody durability) selected for the analysis of humoral responses mounted upon infection. B: Durability of SARS-CoV-2 spike binding antibodies over time. SARS-CoV-2 IgG binding antibody dynamics after infection were described in 137 PARIS participants by an additive mixed model. The early waning period is followed by stabilization. The date of SARS-CoV-2 infection was determined by a positive nucleic acid amplification test, onset of COVID-19 symptoms, or the date of first positive antibody test. Most participants were infected in the first wave of the pandemic. The limit of detection of the SARS-CoV-2 IgG antibody ELISA is set at a titer of 1:80. The blue line represents the mean antibody titer value predicted by the model and the light blue shaded region represents the 95% confidence interval of the mean. C: SARS-CoV-2 antibody durability depends on the initial levels of antibodies. The antibody durability group was split by the antibody titer at enrollment (pink: ≥1:800, N: 81; orange: <1:800, N: 56). Both groups demonstrate broadly similar dynamics, with the early waning period being most evident in the group with higher initial SARS-CoV-2 spike binding antibody titers. Titers equal to/or above 1:800 were defined as high. The pink and orange lines represent the mean antibody titer value predicted by the model and the light pink/orange shaded regions represent the 95% confidence interval of the mean.

To model spike-binding antibody kinetics, we analyzed a total of 813 distinct spike binding measurements from 137 participants (median: five study visits; IQR: 4-8 visits per participant, longitudinal follow-up of two to six months up to 400 days post-infection, see **Table 1, Supplemental Figure 1**). The date of symptom onset or positive NAAT was used as Day 0 when available. Alternatively, we used the date of first positive SARS-CoV-2 antibody assay as Day 0 for 21 participants. Spike binding IgG antibody titers were highly variable among COVID-19 survivors, with titers ranging between 1:80 and 1:6,400. The majority (59.1%) of participants had SARS-CoV-2 binding antibody titers above 1:800 at their baseline visit. We noted that the antibody levels decreased over the first three months, followed by a relative stabilization that persisted up to one year post-infection (**Figure 1B**). Given the large variation in the initial antibody levels, we modeled whether the slopes for those with titers above 1:800 were different from the slopes measured for those with lower antibodies (less than 1:800). SARS-CoV-2 spike binding antibody kinetics between the two groups were comparable, with the initial decay being more pronounced in the high antibody group (**Figure 1C**).

We next tested whether demographic variables such as sex or age were associated with the durability of spike antibody durability by modeling the impact of sex and age on antibody levels over the course of the observation period. We found that more advanced age (e.g., 40 years or older) was associated with 1.62-fold higher antibody levels (95% CI: 1.20-2.19) compared to younger participants. Sex was also associated with the level of SARS-CoV-2 spike antibodies, with antibody levels being 1.40-fold higher in female participants (95% CI: 1.03-1.92) than male participants (**Supplementary Table S1**).

All the participants with documented SARS-CoV-2 infection also mounted detectable antibody responses but we wondered whether seropositive individuals would turn seronegative during the observation period. We found that 6% (8/137) of the initially seropositive participants in the Antibody Durability Dataset tested negative on subsequent visits occurring over up to eleven months of study follow-up. All eight of these participants were initially in the lower baseline antibody group (below 1:800 titer) pointing to a significantly higher risk of seroreversion for individuals with initially lower antibody titers (**Figure 2**, Kaplan-Meier estimate).

**Figure 2:**
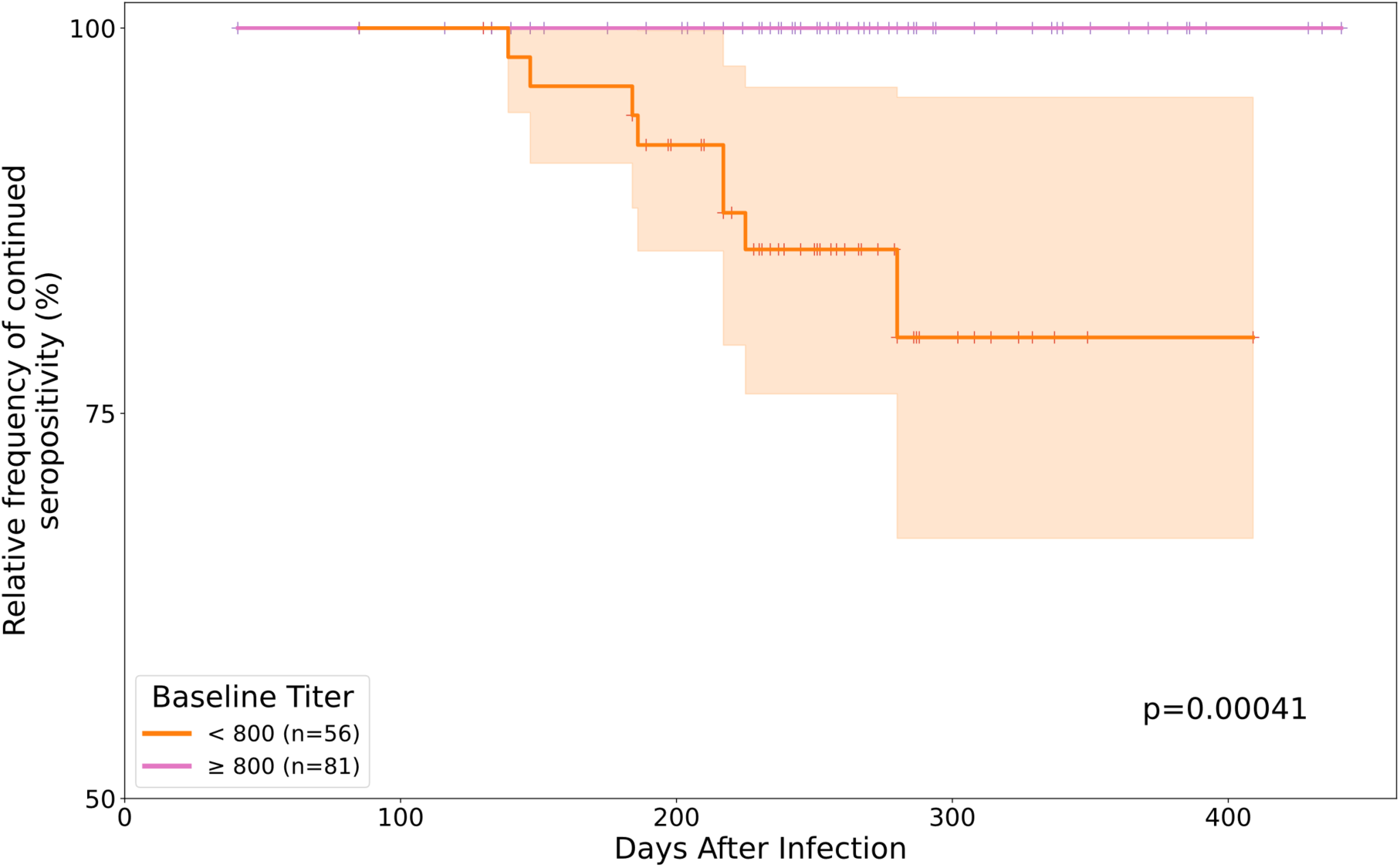
Risk of seroreversion in the antibody durability dataset. The Kaplan-Meier estimate indicates that the risk of seroreversion was significantly higher for the participants with lower antibody titer at the time study enrollment (N=56 <1:800 titer [orange], N=81 >1:800 titer [pink]). Of the 137 participants included in the antibody durability dataset, only eight [6%] participants seroreverted, all of which were initially in the lower antibody titer group.

Finally, we tested whether spike-binding IgG antibodies were associated with protection from re-infection with genetically similar SARS-CoV-2 variants. Between July 2020 and August 2021, we documented a total of 11 new SARS-CoV-2 infections in PARIS participants (**Figure 3A**). Of note, 10/11 of these infections occurred at a time when only ancestral viral variants circulated in the NY metropolitan area (**Figure 3A**). All but one of the SARS-CoV-2 infections occurred in naïve participants. One infection was found in a participant with prior COVID-19, albeit without detectable antibodies at time of re-infection (**Figure 3B**). Thus, detectable spike-binding IgG antibodies mounted upon infection are associated with significant protection from re-infection (Fisher’s Exact Test, p=0.001) in this pre-vaccine and pre-Omicron era of the COVID-19 pandemic.

**Figure 3:**
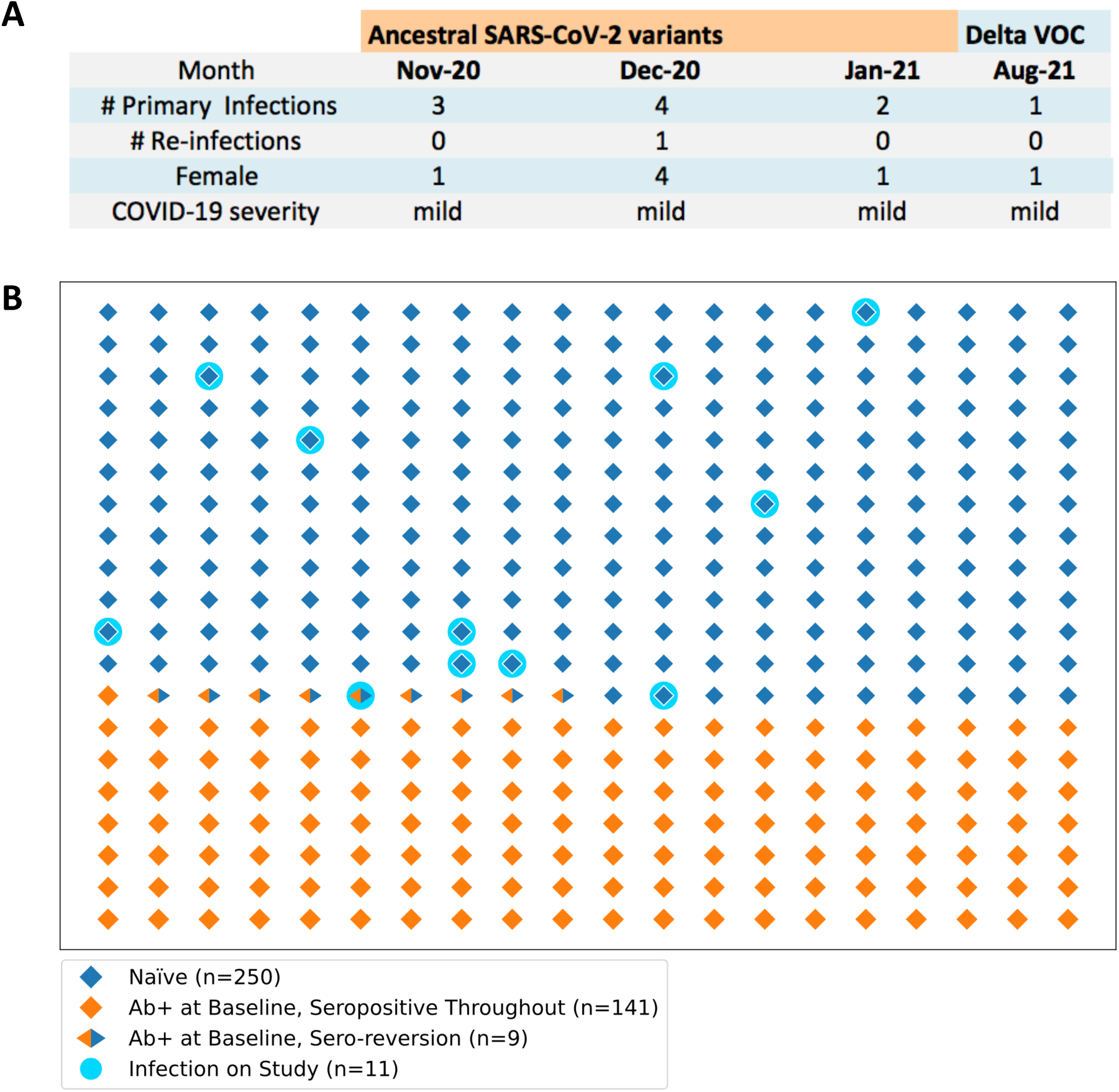
SARS-CoV-2 infections occurring in PARIS participants. A: Summary of the new infections documented in the protection dataset (N=400). The circulating viral variants, the month of infection, the sex of the infected participants and the COVID-19 severity is listed for the 11 SARS-CoV-2 infections documented in the protection dataset between April 2020 and August 2021. B: Graphic representation of the frequency of SARS-CoV-2 infections in the seropositive (N=149, orange diamonds) and seronegative (N=251, blue diamond) study participants. Participants who seroreverted are indicated by the orange/blue symbol. On-study infections occurred in 11 study participants without detectable SARS-CoV-2 spike binding antibodies (turquoise circles). 10/11 were naïve (blue diamond symbols) and only one participant had a documented prior COVID-19 but no antibodies at the time of infection.

## Discussion

Several studies have evaluated the durability of serum SARS-CoV-2 IgG antibodies (14). While immune responses to SARS-CoV-2 infection and vaccination and their respective protective effects have been analyzed at rapid speed and in much detail, many open questions remain. We leveraged the fact that most seropositive PARIS participants were infected in the first pandemic wave (March-April 2020) with very homogenous SARS-CoV-2 strains (15). An additional strength of the PARIS cohort is the frequent, longitudinal sample and data collection (every 2-4 weeks), which allows for a high level of granularity in the modeling of the durability and effectiveness of SARS-CoV-2 antibody responses. We first evaluated the stability of spike-binding IgG antibody titers over time. Typical antibody responses to infection are characterized by an initial strong peak, driven by short-lived plasmablasts in the peripheral blood circulation, followed by a decline and an eventual stabilization at a level of antibody that is produced by long-lived plasmablasts in the bone marrow (16-18). This was exactly the pattern that we observed in our analysis: high SARS-CoV-2 antibody titers declined initially but stabilized over the following months. Overall, seroreversion was rare with only 6% of the COVID-19 survivors having antibody levels wane to below the level of detection of our sensitive full-length spike IG binding antibody ELISA. Interestingly, there were large differences in the spike-binding antibody titers across participants with those 40 years and older having higher antibody titers compared to younger individuals. This phenomenon has been observed before (19, 20). Our observation that female participants have higher antibody titers than male participants stands in contrast to several previous studies reporting that males have higher SARS-CoV-2 antibody levels (4). The PARIS cohort comprises mostly younger and overall healthy health care workers with almost exclusively mild infections, so it is conceivable that this sex difference becomes less apparent when more severe COVID-19 manifestations known to result in higher antibody levels (20) are included in the analysis. Of note other than the magnitude, there was no sizable difference in the kinetics of antibody levels depending on age or sex. Additional studies in longitudinal cohorts with as frequent sampling as done in the PARIS cohort are needed to independently replicate our observations.

Several studies from the pre- (20-26) and post-Delta (B.1.617.2) (27, 28) era suggest that protection from reinfection ranges around 80-90% if the circulating SARS-CoV-2 variants are antigenically similar to the ones responsible for the initial infections. Only the appearance of Omicron has led to an increase of reinfections (28, 29). Our cohort study supports this notion since we did not document any reinfections in study participants who were previously infected and maintained detectable levels of spike binding antibodies. Indeed, ten naïve individuals and one individual with an initially low titer who sero-reverted prior to re-infection were infected during the observation period. These findings suggest significant protection from reinfection and hint at the importance of the level of spike-binding antibody titers in protection. Of note, the presence of spike-binding antibodies was also correlated with protection in several other studies (21, 30-32). Antibody titers against the receptor binding domain and the full-length spike, as well as neutralizing antibodies, have recently been proposed as correlates of protection of vaccine induced immunity (33-35). Importantly, the data in these studies were generated before the Omicron variants started to circulate at larger scale in our community. Similarly, the current analysis was conducted prior to the circulation of SARS-CoV-2 variants of concern in the NYC metropolitan area. Our data underscores that spike binding antibodies protect against infection with antigenically similar viral strains. Protection against heterologous, antigenically distinct variants such as Omicron is likely limited based on the pronounced reduction in virus neutralization (36-39).

This study has several strengths. First, the prospective nature of the cohort allowed us to assess how antibody responses against SARS-CoV-2 following natural infection changed over time. Second, the repeated sampling/testing of this study provides a perspective on a much more granular scale than previous analyses. Finally, this study is based on data collected prior to the introduction of SARS-CoV-2 vaccines and the wide circulation of variants of concern that are highly antigenically distinct (e.g., Omicron). As such, it provides a useful baseline against which newer data can be compared to answer important questions regarding the relative severity of new variants, the strength and durability of antibody responses against SARS-CoV-2 variants, and the impact of immune histories on the breadth of immune responses.

This analysis did, however, also have a few limitations. First, since we started enrollment during the first wave, a good portion of participants were unable to get molecular tests at the time of infection and we relied on retrospective reports of clinical signs and symptoms suggestive of COVID-19 for illness onset date. As such, recall bias in reported illness onset is a possibility. However, we anticipate that this exerted only a minor impact on our conclusions given the relatively homogenous exposures of participants who are all health care workers. Second, with healthcare worker vaccination beginning in December 2020 we were unable to effectively assess how circulating variants of concern may affect one’s risk of re-infection following natural infection. The increase in vaccinated participants (excluded from this analysis), while fortunate, also resulted in a smaller sample size at the end of the follow-up period extending into August 2021.

In conclusion, our study shows that SARS-CoV-2 infection provides strong protection from reinfection and this protection may be associated with the presence of spike binding antibodies. In addition, it suggests that antibody levels induced by infection with ancestral SARS-CoV-2 variants are relatively stable over time and that the rate of seroreversion is low when measuring SARS-CoV-2 spike binding IgG antibodies.

## Data Availability

All data produced in the present study are available upon reasonable request to the authors.

## Acknowledgements

We thank the study participants for their generosity and willingness to participate in longitudinal COVID-19 research studies. None of this work would be possible without their contributions.

We very appreciative of the support of the Mount Sinai’s leadership throughout the COVID-19 pandemic. We want to especially thank Drs. Peter Palese, Carlos Cordon-Cardo, Dennis Charney, David Reich and Kenneth Davis. We would also like to thank Daniel Caughey for expert administrative assistance and Dr. Andrew Brouwer for his thoughtful comments and suggestions. This work is part of the PARIS/SPARTA studies funded by the NIAID Collaborative Influenza Vaccine Innovation Centers (CIVIC) contract 75N93019C00051. In addition, this work was also partially funded by the NIAID Centers of Excellence for Influenza Research and Response (CEIRR) contract 75N93021C00014 as well as by anonymous donors. This work is part of the NIAID SARS-CoV-2 Assessment of Viral Evolution (SAVE) program.

## Supplemental Data

**Supplemental Table S1:**
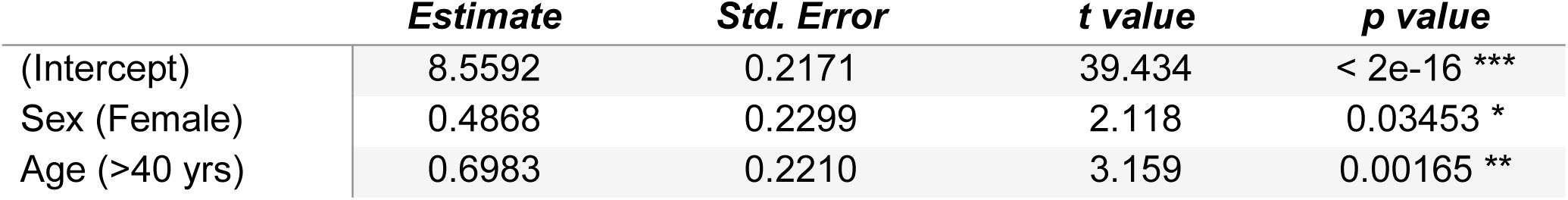
Additive mixed model results regarding variables influencing the level of SARS-CoV-2 spike binding IgG antibodies. The model indicates significant effects on SARS-CoV-2 antibody levels due to both age (>40 years of age) and sex (female). Significance was set at p < 0.05. Estimates are on a log 2 scale.

|  | *Estimate* | *Std. Error* | *t value* | *p value* |
| --- | --- | --- | --- | --- |
| (Intercept) | 8.5592 | 0.2171 | 39.434 | < 2e-16 *** |
| Sex (Female) | 0.4868 | 0.2299 | 2.118 | 0.03453 * |
| Age (>40 yrs) | 0.6983 | 0.2210 | 3.159 | 0.00165 ** |

## Supplemental Figure legends

**Supplemental Figure S1:**
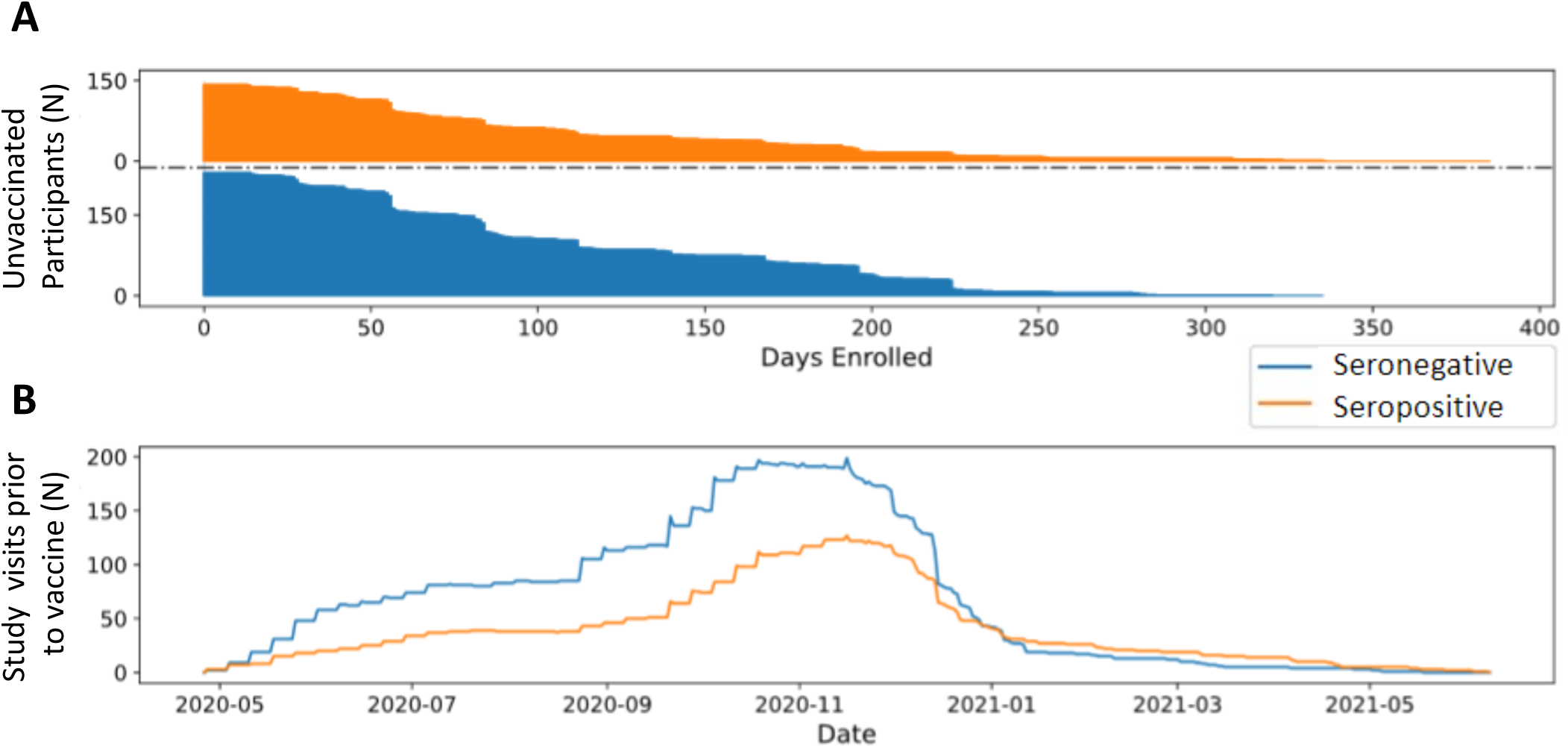
Duration of study follow-up prior for unvaccinated PARIS participants included in the protection dataset. (A) The time between first and final pre-vaccine antibody measurements for each participant is shown graphically. The total length of follow-up for unvaccinated participants varies substantially since the SARS-CoV-2 vaccine rollout for health care workers started in the middle of December 2020. (B) Enrollment in the PARIS cohort and follow up are shown. PARIS began in April 2020 and continued recruitment throughout the year. The graph summarizes all the pre-vaccination visits for each participant. The number of unvaccinated participants in PARIS declined sharply when SARS-CoV-2 vaccinations became available in December 2020 limiting the number of follow up visits included in the study. Vaccination became mandatory for Mount Sinai staff members in September 2021.

## References

1. Huang C, Wang Y, Li X, Ren L, Zhao J, Hu Y, et al. Clinical features of patients infected with 2019 novel coronavirus in Wuhan, China. Lancet. 2020;395(10223):497–506.

2. Zhou P, Yang XL, Wang XG, Hu B, Zhang L, Zhang W, et al. A pneumonia outbreak associated with a new coronavirus of probable bat origin. Nature. 2020.

3. Fafi-Kremer S, Bruel T, Madec Y, Grant R, Tondeur L, Grzelak L, et al. Serologic responses to SARS-CoV-2 infection among hospital staff with mild disease in eastern France. EBioMedicine. 2020;59:102915.

4. Wajnberg A, Mansour M, Leven E, Bouvier NM, Patel G, Firpo-Betancourt A, et al. Humoral response and PCR positivity in patients with COVID-19 in the New York City region, USA: an observational study. Lancet Microbe. 2020;1(7):e283–e9.

5. Wajnberg A, Amanat F, Firpo A, Altman DR, Bailey MJ, Mansour M, et al. Robust neutralizing antibodies to SARS-CoV-2 infection persist for months. Science. 2020;370(6521):1227–30.

6. Long QX, Tang XJ, Shi QL, Li Q, Deng HJ, Yuan J, et al. Clinical and immunological assessment of asymptomatic SARS-CoV-2 infections. Nat Med. 2020.

7. Dan JM, Mateus J, Kato Y, Hastie KM, Yu ED, Faliti CE, et al. Immunological memory to SARS-CoV-2 assessed for up to 8 months after infection. Science. 2021;371(6529).

8. Grandjean L, Saso A, Ortiz AT, Lam T, Hatcher J, Thistlethwayte R, et al. Long-Term Persistence of Spike Antibody and Predictive Modeling of Antibody Dynamics Following Infection with SARS-CoV-2. Clin Infect Dis. 2021.

9. Carreño JM, Mendu DR, Simon V, Shariff MA, Singh G, Menon V, et al. Longitudinal analysis of severe acute respiratory syndrome coronavirus 2 seroprevalence using multiple serology platforms. iScience. 2021;24(9):102937.

10. Hernandez MM, Gonzalez-Reiche AS, Alshammary H, Fabre S, Khan Z, van De Guchte A, et al. Molecular evidence of SARS-CoV-2 in New York before the first pandemic wave. Nat Commun. 2021;12(1):3463.

11. Stadlbauer D, Tan J, Jiang K, Hernandez MM, Fabre S, Amanat F, et al. Repeated cross-sectional sero-monitoring of SARS-CoV-2 in New York City. Nature. 2021;590(7844):146–50.

12. Yang W, Kandula S, Huynh M, Greene SK, Van Wye G, Li W, et al. Estimating the infection-fatality risk of SARS-CoV-2 in New York City during the spring 2020 pandemic wave: a model-based analysis. Lancet Infect Dis. 2021;21(2):203–12.

13. Amanat F, Stadlbauer D, Strohmeier S, Nguyen THO, Chromikova V, McMahon M, et al. A serological assay to detect SARS-CoV-2 seroconversion in humans. Nat Med. 2020;26(7):1033–6.

14. Egbert ER, Xiao S, Colantuoni E, Caturegli P, Gadala A, Milstone AM, et al. Durability of Spike Immunoglobin G Antibodies to SARS-CoV-2 Among Health Care Workers With Prior Infection. JAMA Netw Open. 2021;4(8):e2123256.

15. Gonzalez-Reiche AS, Hernandez MM, Sullivan MJ, Ciferri B, Alshammary H, Obla A, et al. Introductions and early spread of SARS-CoV-2 in the New York City area. Science. 2020;369(6501):297–301.

16. Laidlaw BJ, Ellebedy AH. The germinal centre B cell response to SARS-CoV-2. Nat Rev Immunol. 2022;22(1):7–18.

17. Turner JS, Kim W, Kalaidina E, Goss CW, Rauseo AM, Schmitz AJ, et al. SARS-CoV-2 infection induces long-lived bone marrow plasma cells in humans. Nature. 2021.

18. Wrammert J, Smith K, Miller J, Langley WA, Kokko K, Larsen C, et al. Rapid cloning of high-affinity human monoclonal antibodies against influenza virus. Nature. 2008;453(7195):667–71.

19. Bag Soytas R, Cengiz M, Islamoglu MS, Uysal BB, Ikitimur H, Yavuzer H, et al. Does the COVID-19 seroconversion in older adults resemble the young? J Med Virol. 2021;93(10):5777–82.

20. Maier HE, Kuan G, Saborio S, Carrillo FAB, Plazaola M, Barilla C, et al. Clinical Spectrum of Severe Acute Respiratory Syndrome Coronavirus 2 Infection and Protection From Symptomatic Reinfection. Clinical Infectious Diseases. 2021.

21. Hall VJ, Foulkes S, Charlett A, Atti A, Monk EJM, Simmons R, et al. SARS-CoV-2 infection rates of antibody-positive compared with antibody-negative health-care workers in England: a large, multicentre, prospective cohort study (SIREN). Lancet. 2021;397(10283):1459–69.

22. Pilz S, Chakeri A, Ioannidis JP, Richter L, Theiler-Schwetz V, Trummer C, et al. SARS-CoV-2 re-infection risk in Austria. Eur J Clin Invest. 2021;51(4):e13520.

23. Lumley SF, O’Donnell D, Stoesser NE, Matthews PC, Howarth A, Hatch SB, et al. Antibody Status and Incidence of SARS-CoV-2 Infection in Health Care Workers. N Engl J Med. 2020.

24. Sheehan MM, Reddy AJ, Rothberg MB. Reinfection Rates Among Patients Who Previously Tested Positive for Coronavirus Disease 2019: A Retrospective Cohort Study. Clin Infect Dis. 2021;73(10):1882–6.

25. Hansen CH, Michlmayr D, Gubbels SM, Mølbak K, Ethelberg S. Assessment of protection against reinfection with SARS-CoV-2 among 4 million PCR-tested individuals in Denmark in 2020: a population-level observational study. Lancet. 2021.

26. Harvey RA, Rassen JA, Kabelac CA, Turenne W, Leonard S, Klesh R, et al. Association of SARS-CoV-2 Seropositive Antibody Test With Risk of Future Infection. JAMA Intern Med. 2021;181(5):672–9.

27. Pouwels KB, Pritchard E, Matthews PC, Stoesser N, Eyre DW, Vihta KD, et al. Effect of Delta variant on viral burden and vaccine effectiveness against new SARS-CoV-2 infections in the UK. Nat Med. 2021;27(12):2127–35.

28. Pulliam JRC, van Schalkwyk C, Govender N, von Gottberg A, Cohen C, Groome MJ, et al. Increased risk of SARS-CoV-2 reinfection associated with emergence of the Omicron variant in South Africa. medRxiv. 2021:2021.11.11.21266068.

29. Altarawneh HN, Chemaitelly H, Hasan MR, Ayoub HH, Qassim S, AlMukdad S, et al. Protection against the Omicron Variant from Previous SARS-CoV-2 Infection. N Engl J Med. 2022.

30. Krammer F. Correlates of protection from SARS-CoV-2 infection. Lancet. 2021;397(10283):1421–3.

31. Letizia AG, Ge Y, Vangeti S, Goforth C, Weir DL, Kuzmina NA, et al. SARS-CoV-2 seropositivity and subsequent infection risk in healthy young adults: a prospective cohort study. Lancet Respir Med. 2021.

32. Addetia A, Crawford KHD, Dingens A, Zhu H, Roychoudhury P, Huang ML, et al. Neutralizing antibodies correlate with protection from SARS-CoV-2 in humans during a fishery vessel outbreak with high attack rate. J Clin Microbiol. 2020.

33. Khoury DS, Cromer D, Reynaldi A, Schlub TE, Wheatley AK, Juno JA, et al. Neutralizing antibody levels are highly predictive of immune protection from symptomatic SARS-CoV-2 infection. Nat Med. 2021.

34. Goldblatt D, Fiore-Gartland A, Johnson M, Hunt A, Bengt C, Zavadska D, et al. Towards a population-based threshold of protection for COVID-19 vaccines. Vaccine. 2022;40(2):306–15.

35. Cromer D, Steain M, Reynaldi A, Schlub TE, Wheatley AK, Juno JA, et al. Neutralising antibody titres as predictors of protection against SARS-CoV-2 variants and the impact of boosting: a meta-analysis. Lancet Microbe. 2022;3(1):e52–e61.

36. Cameroni E, Bowen JE, Rosen LE, Saliba C, Zepeda SK, Culap K, et al. Broadly neutralizing antibodies overcome SARS-CoV-2 Omicron antigenic shift. Nature. 2021.

37. Juan Manuel Carreño, Hala Alshammary, Johnstone Tcheou, Gagandeep Singh, Ariel Raskin, Hisaaki Kawabata, et al. Activity of convalescent and vaccine serum against SARS-CoV-2 Omicron. Nature. 2021;doi: https://doi.org/10.1038/d41586-021-03846-z.

38. Dejnirattisai W, Huo J, Zhou D, Zahradník J, Supasa P, Liu C, et al. SARS-CoV-2 Omicron-B.1.1.529 leads to widespread escape from neutralizing antibody responses. Cell. 2022;185(3):467–84.e15.

39. Muik A, Lui BG, Wallisch AK, Bacher M, Mühl J, Reinholz J, et al. Neutralization of SARS-CoV-2 Omicron by BNT162b2 mRNA vaccine-elicited human sera. Science. 2022;375(6581):678–80.

